# Variability in sensory processing and evoked potentials in Rett syndrome

**DOI:** 10.64898/2026.07.07.26357400

**Authors:** Devorah Kranz, Klara Szilagyi, Karen N Sabol, David Lieberman, Charles Nelson, April R Levin, Michela Fagiolini

## Abstract

**Background:** Rett syndrome (RTT), a rare neurodevelopmental disorder caused primarily by pathogenic variants in the *MECP2* gene, is characterized by severe cognitive, motor, and autonomic impairments. Atypical sensory processing—including co-occurring hypo- and hyper-responsivity—is a core yet poorly understood feature. While evoked potentials (EPs) show delayed and attenuated sensory responses in RTT, the underlying mechanisms of these impairments remain unclear. Inter-trial phase coherence (ITPC), which quantifies trial-by-trial neural response consistency, offers a promising functional biomarker of variability in sensory processing.

**Methods:** We characterized caregiver-reported sensory responsivity in 32 individuals with RTT (all female) and 28 typically developing controls (26 female, 2 male). EPs were then recorded during passive visual and auditory stimulation and ITPC was computed to assess whether variability in the timing of neural responses could account for reduced EP amplitudes and atypical sensory responsivity.

**Results:** Hypo- and hyper-responsivity to sensory stimuli were both significantly elevated in RTT and were positively correlated, co-occurring within individuals. ITPC was significantly reduced in RTT across visual and auditory modalities and was associated with reduced EP amplitudes. Notably, reduced ITPC in visual-evoked potentials was further associated with elevated visual responsivity and greater behavioral symptom severity.

**Conclusions:** Increased variability in neural response timing may contribute to both reduced EPs and atypical sensory responsivity in RTT, supporting ITPC as a functional biomarker. Decreased temporal precision of neural activity may explain the co-occurrence of hypo- and hyper-responsivity and provide a unifying framework for sensory dysfunction across neurodevelopmental disorders.

## Introduction

Rett syndrome (RTT) is a rare neurodevelopmental disorder affecting about 1 in 10,000 individuals, predominantly caused by *de novo* pathogenic variants in the X-linked *MECP2* gene.(1) These mutations are also associated with an increased likelihood of autism spectrum disorder (ASD); RTT is often used as a model for ASD because many symptoms overlap, although ASD can be hard to diagnose in RTT given cognitive and motor challenges.(2) Primarily affecting females, individuals with RTT initially experience a typical period of development followed by rapid and regressive loss of purposeful hand use and verbal language. Cognitive, motor, and autonomic dysfunctions including epilepsy, irregular or absent gait, and breathing irregularities are additional common symptoms.(3,4) More recently, differences in sensory processing—a key feature of many neurodevelopmental disorders (NDDs), especially ASD(5–7)—have also been identified as a core feature of RTT.(8,9)

Atypical sensory processing can include hypo-responsivity (reduced behavioral responses to sensory stimuli, sometimes with compensatory sensory seeking behaviors) and hyper-responsivity (exaggerated or avoidant behavioral responses to sensory stimuli), with cascading consequences impacting social interactions, cognitive functioning, and quality of life.(10–13) In ASD, both hypo- and hyper-responsivity have been documented in visual, auditory, and tactile modalities,(5,6,14,15) but are generally conceptualized as separate, distinct constructs, representing opposing ends of a sensory processing spectrum. However, their frequent co-occurrence within the same individual(15–17) suggests they may instead reflect dynamic, moment-to-moment variability in sensory processing rather than fixed or opposing profiles.(18) In RTT, sensory hyper-responsivity, hypo-responsivity, and seeking behaviors have similarly been reported,(19) yet objective biomarkers that directly capture the relationship between these various aspects of sensory processing and their underlying neural dynamics remain limited.

Electroencephalography (EEG), and particularly evoked potentials (EPs)—offer powerful tools to investigate the neurophysiological underpinnings of atypical sensory processing in RTT.(20–23) EPs capture the time course of neural responses to sensory stimuli and can be reliably elicited through passive paradigms requiring no behavioral response, making them well-suited for studying individuals with RTT and other NDDs. EP abnormalities have been shown to correlate with cognitive(24) and clinical outcomes,(20,21) and have been well-documented across NDDs with strong features of sensory processing abnormalities and some overlapping features with RTT including ASD,(25–27) Down Syndrome,(28) Prader-Willi syndrome,(29) Fragile X syndrome,(30,31) Angelman Syndrome,(32) Phelan McDermid syndrome,(33) CDKL5 deficiency disorder(34) and others.

Auditory evoked potentials (AEPs) have consistently revealed abnormalities in RTT,(8,21,22,35– 39) most prominently characterized by the altered amplitude of components such as P1-N1 and N1-P2 which associate with clinical severity.(8,21,37) Studies of higher-order auditory processing assessed through oddball paradigms also reveal delayed and prolonged responses in RTT,(22,35,37) and individuals with RTT show impaired auditory discrimination abilities, particularly in response to stimulus duration changes, suggesting disrupted auditory memory span.(8,40) Visual evoked potentials (VEPs) also exhibit substantial abnormalities in RTT, with significant differences in P1 amplitude and N2 latency observed in individuals with RTT and altered VEP waveforms in *Mecp2* heterozygote female mice, both correlating with disease severity and progression.(20,21) Notably, preclinical studies in Mecp2-deficient mouse models have revealed mechanisms that may underlie disrupted visual cortical processing, pointing to reduced neural activity,(41) impaired critical period plasticity,(42) and progressive thalamocortical disruption.(43) However, despite extensive evidence of impaired EPs across sensory modalities in RTT, the mechanisms underlying disruptions to these neural responses remain incompletely understood.

EPs are traditionally resolved by averaging across many trials, an approach that assumes consistent, phase-locked neural responses to repeated presentations of identical stimuli within individuals.(44,45) Indeed, EP components in typically developing (TD) individuals emerge largely from consistent phase alignment of neural oscillations across trials.(46–49) However, while this assumption may hold in neurotypical populations, it may not accurately reflect underlying neural dynamics in NDDs with variable sensory responsivity; if responses vary in their temporal alignment across trials, averaging may artificially attenuate the EP amplitude rather than reflect a true reduction in response magnitude, and critical information contained within intra-individual variability may be lost.(50) Such trial-by-trial variability can thus provide a potential mechanism for the attenuated EPs observed in RTT.

Time-frequency analyses, and in particular inter-trial phase coherence (ITPC), provides a quantitative measure of the consistency of the neural response by assessing trial-by-trial phase-locking. ITPC reflects the extent to which neural responses occur at a similar phase across trials relative to stimulus onset, ranging from random phase distribution (low ITPC) to perfect phase synchronization (high ITPC). ITPC captures the temporal precision of neural responses rather than their amplitude per se. Reduced ITPC has been demonstrated in ASD,(51–55) and recent work in RTT has identified elevated neural variability in auditory responses, including ITPC.(38) However, ITPC in visual-evoked potentials has not yet been studied in RTT, and it remains unclear whether neural response variability relates to behavioral measures of sensory processing.

Here, we tested three main hypotheses: (1) that individuals with RTT would exhibit co-occurring hypo- and hyper-responsivity to sensory stimuli, (2) that individuals with RTT would show reduced ITPC in both AEP and VEP, reflecting increased within-individual variability in the timing of neural responses across trials, and (3) that this neural variability would be associated with both reduced EP amplitude and altered sensory responsivity.

## Methods and Materials

### Participants

Data were acquired at BCH as part of the Rett Syndrome Biomarkers Study (RSBS), through the Intellectual and Developmental Disabilities Research Center (IDDRC) at Boston Children’s Hospital. The experimental protocol was approved and overseen by the Institutional Review Board at Boston Children’s Hospital.

Participants included individuals with Rett Syndrome (RTT: n=32, 1-45 years old) and typically developing controls (TD: n=28, 2-23 years old). While many prior studies in RTT have focused on a narrower age range, we chose this wider range to capture the full spectrum of the disorder; age effects were evaluated and accounted for in analyses as described below. Participants were recruited through hospital-based Rett clinics, a research registry, clinical referrals, community sources, and word of mouth. Informed consent was obtained from all guardians or TD participants ≥18 years old, and assent was obtained from all participants developmentally able.

RTT inclusion criteria included age 12 months to 50 years and a diagnosis of RTT with a pathogenic or likely pathogenic variant in the *MECP2* gene. TD participants were approximately age and sex-matched, with no known genetic diagnoses or conditions associated with ASD or intellectual disability. The primary language of all participants was English. RTT exclusion criteria included participation in an investigational drug study within 30 days prior to enrollment.

Demographic and other characteristics are reported in Table 1.

**Table 1.** Participant Information.

|  | Diagnostic Group |  |
| --- | --- | --- |
|  | RTT (n = 32) | TD (n = 28) |
| **Clinical Severity Score** |  |  |
| <10 | 3 |  |
| 10->19 | 16 |  |
| 20->29 | 11 |  |
| 30->39 | 2 |  |
| **Clinical Global Impressions of Severity** |  |  |
| <4 | 1 |  |
| 4 | 13 |  |
| 5 | 17 |  |
| >5 | 1 |  |
| **Rett Diagnosis** |  |  |
| Typical | 20 |  |
| Atypical | 12 |  |
| **Seizure History** |  |  |
| No | 11 |  |
| Yes | 21 |  |
| **Sex** |  |  |
| Female | 32 | 26 |
| Male | 0 | 2 |
| **Age** |  |  |
|  | Range: 1-45 | Range: 2-23 |
|  | Median: 9 | Median: 7 |
|  | IQR: 16.25 | IQR: 6.25 |
| **Usable VEP** |  |  |
| Yes | 31 | 28 |
| No | 1 | 0 |
| **Usable AEP** |  |  |
| Yes | 27 | 26 |
| No | 0 | 0 |

### Phenotypic and clinical measures: sensory processing

Parents or caregivers completed detailed medical history questionnaires and phenotypic assessments during the visit or remotely within 6 months of the visit date. Sensory processing phenotype was assessed using the Child Sensory Profile 2 (SP2), a standardized caregiver-report questionnaire that measures sensory processing patterns in everyday life.(5,56) We used the Child SP2 for all participants regardless of chronological age. Although validated for individuals up to 14 years of age, the Child SP2 was used outside this normative range to better reflect the developmental level of RTT participants. For each participant, overall hyper-responsivity scores were derived by summing Sensitivity and Avoiding quadrants, and hypo-responsivity scores by summing Registration and Seeking quadrants.(15) Visual and auditory responsivity sub-scores were also analyzed. Higher scores indicate greater sensory processing difficulties. We use the term “responsivity” for these measures throughout, consistent with the taxonomy proposed by He, et al., 2023.(13) The SP2 was introduced later in the study, so not all participants with EP data had SP2 data. SP2 analyses included 29 RTT and 25 TD individuals; among these, VEP data were available for 24 RTT and 25 TD, and AEP data for 24 RTT and 23 TD participants.

### Phenotypic and clinical measures: clinical severity

Clinical severity in RTT was assessed by clinicians using the Clinical Severity Scale (CSS)(57) and Clinical Global Impressions of Severity (CGI-S).(58,59) Scores were typically obtained on the same day as EP recordings (when not available on the same day, scores from the closest clinical assessment within 6 months were used). Clinicians also categorized RTT diagnosis as typical or atypical and recorded seizure frequency. Clinical measures are summarized in Table 1.

Behavioral symptom severity was assessed using the Rett Syndrome Behavior Questionnaire (RSBQ),(60–62) a caregiver-report measure of neurobehavioral symptoms increasingly used in clinical and research settings.(63–67) Higher scores (maximum = 90) reflect greater symptom severity and frequency of behavioral abnormalities.

### EEG data collection

Participants were seated 65 cm from a monitor in a dimly lit, sound-attenuated, electrically shielded room. Participants sat independently or in a caregiver’s lap; caregivers were instructed not to converse with the participant during data collection. A behavioral assistant was present to maintain engagement and redirect attention when needed. Continuous EEG data were recorded using a Net Amps 400 amplifier with a 128-channel Hydrocel Geodesic Net (Magstim Electrical Geodesic, Inc., Eugene, OR). Data were sampled at 1,000 Hz with reference to the electrode “Cz”. Stimuli were presented using Presentation software with the following parameters:

#### VEP

400 trials of a reversing black-and-white checkerboard stimulus (0.5 cycles/degree), presented at a reversal rate of 2 Hz (500 ms per trial).(20,21) Stimulus presentation was contingent on visual attention: eye gaze was monitored and stimulus delivery was paused if gaze deviated from the screen.

#### AEP

250 trials of 500 Hz sinusoidal tones (300 ms duration) presented at 60dB.(21) Interstimulus intervals were jittered between 1-3 seconds. A silent, greyscale movie (typically *WALL-E)* was presented on a separate screen to maintain engagement. This is standard procedure for obtaining high-quality EEG and EPs in individuals with NDDs to reduce movement and maintain engagement without introducing time-locked sensory confounds.(30,68)

### EEG data pre-processing

EEG preprocessing was conducted using the Batch EEG Automated Processing Platform (BEAPP), allowing consistent and standardized preprocessing across EEG data from all participants.(69) Within BEAPP, artifact detection and correction were performed using the Harvard Automated Processing Pipeline for Electroencephalography (HAPPE), a MATLAB-based EEG processing pipeline optimized for data collected from young children and/or study participants with NDDs.(70) Data were bandpass filtered (1-30 Hz) and resampled to 250 Hz. Line noise at 60 Hz was removed using CleanLine’s multi-taper method. Per standard HAPPE parameters, artifact including movement, muscle, and eye blinks were detected and removed using wavelet thresholding. Bad channels were interpolated using spherical interpolation and data were re-referenced to the average reference. Data were segmented into epochs from −400 ms to +500 ms relative to stimulus onset. Epochs were rejected based on HAPPE’s recommended amplitude thresholds (±40μV) and joint probability criteria. For each participant, 185 (VEP) or 200 (AEP) artifact-free epochs were randomly selected for further analysis.

### EEG rejection criteria

Participants were excluded if they had insufficient EEG data or if they exceeded ±3 standard deviations on HAPPE quality metrics, including number of good channels, retained artifact probability (mean and median), percent of independent components rejected, and percent of signal variance retained after artifact removal. Based on these criteria, 1 participant was excluded from VEP analysis, and none from AEP analysis (Table 1). Note that the AEP paradigm was introduced later in the study.

### Evoked potentials

For each participant, evoked potentials were computed by averaging across trials, and component amplitudes were defined as the peak voltage within a predefined time window (“P and “N” being the highest or lowest components).

#### VEP

Analyses were conducted at the occipital electrode Oz, corresponding to maximal visual response. Topographic maps were used to confirm that Oz captured the peak voltage response across participants. The window for the early negative component (N1) was defined as 50-75ms, and for the positive component (P1) as 75-125ms.

#### AEP

Analyses were conducted at the fronto-central electrode Fz. Topographic maps were used to confirm that Fz captured the peak voltage response across participants. The window for the P1 component was defined as 75-125ms, and for the N1 component as 125-175ms.

### Inter-trial phase coherence (ITPC)

Time frequency decomposition was performed using Morlet wavelet convolution. ITPC was calculated for each participant as the circular coherence (mean resultant vector length) across phase values extracted at each time-frequency point, where the length of the average vector indicates the degree of coherence and its direction indicates the dominant phase.(71) The formula is given as:

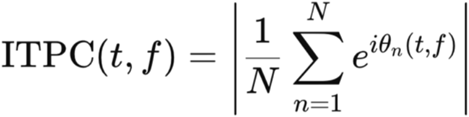

Where θ*n*(*t*,f) represents the phase angle at a given trial *n*, time *t*, and frequency *f*. ITPC values range from 0 (random phase distribution) to 1 (perfect phase alignment).

To visualize the difference between groups, ITPC was averaged across participants and plotted as time-frequency maps with the analyzed frequency range spanning 8-30 Hz. For statistical analysis, ITPC was averaged across frequencies within predefined time windows corresponding to EP components, yielding a single value per participant per component (e.g. VEP P1).(31,38)

### Statistical analyses

Independent-samples t-tests were used to compare phenotypic, EP, and ITPC measures between groups. Welch’s correction was applied when sample size or variance varied widely. Within group comparisons were assessed using paired t-tests. Associations between variables were evaluated using linear regression analyses separately within RTT and TD groups; False Discovery Rate (FDR) correction was applied to account for multiple comparisons. All analyses were performed in MATLAB.

## Results

### Atypical responsivity to sensory stimuli in RTT

We first evaluated sensory processing phenotype in Rett syndrome using the SP2, which measures sensory processing patterns in everyday life. TD participants typically fell within the expected standardized range (40-60) for both hypo- and hyper-responsivity(5) (Fig. 1A-B). The RTT group exhibited significantly higher scores compared to TD, indicating greater sensory challenges in both hypo-responsivity (range: 43-168) and hyper-responsivity (range: 44-115) (P < .001 for both comparisons; Fig. 1A-B).

**Figure 1.**
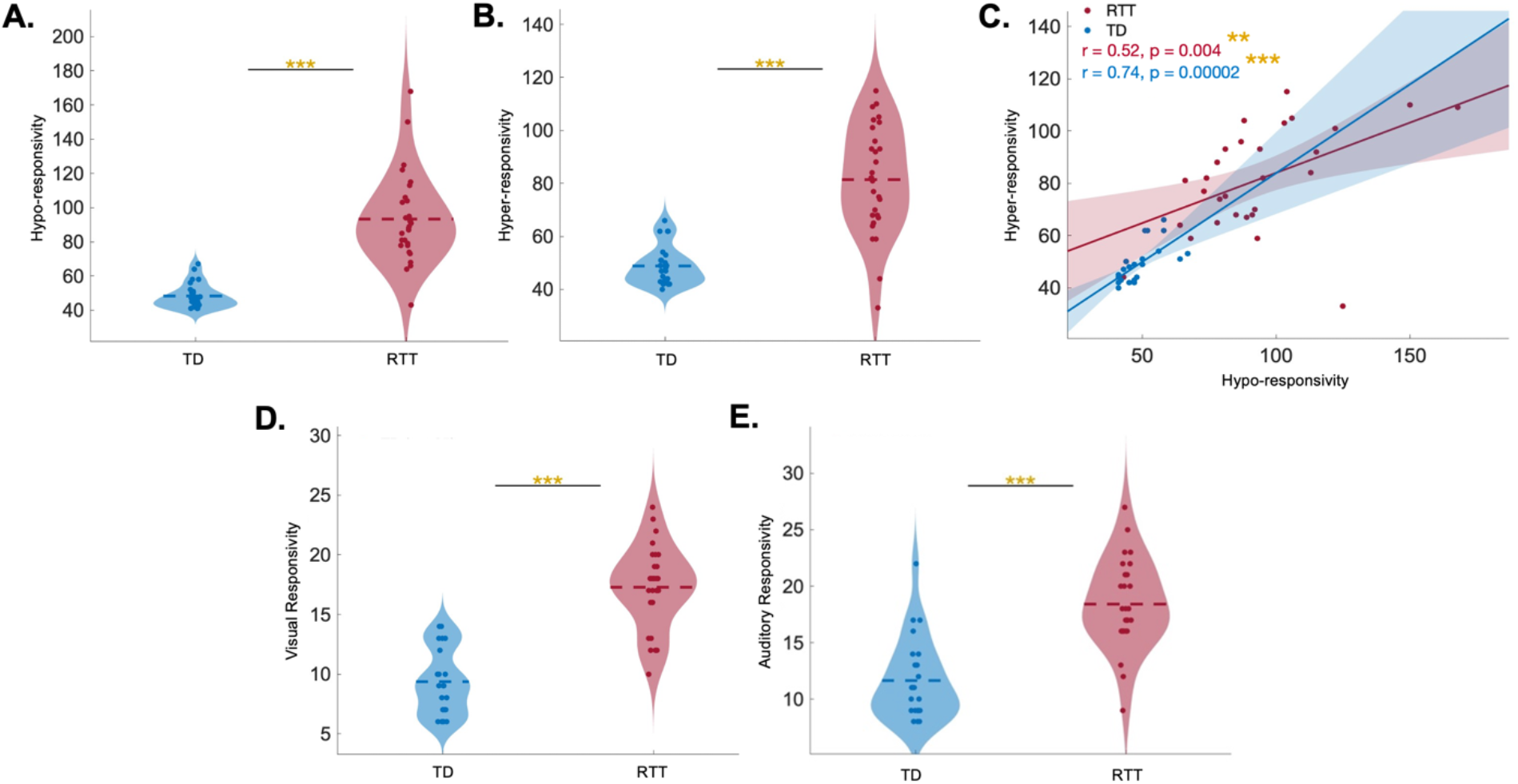
Atypical responsivity to sensory stimuli in RTT. Circles represent individual participant scores on the Sensory Profile 2. TD participants (n=25) are shaded in blue and RTT participants (n=29) are shaded in red. Statistical significance is indicated as follows: *** indicates P<0.001, ** indicates P<0.01, and * indicates P<0.05. **(A, B)** Violin plots of overall hypo- and hyper-responsivity scores for each group (hypo-responsivity in A; hyper-responsivity in B). Violin widths represent kernel density estimates; dashed horizontal lines indicate group means. **(C)** Association between overall hypo- and hyper-responsivity for each group. Shaded areas indicate 95% confidence intervals around linear regression lines. **(D, E)** Violin plots of sensory modality–specific responsivity scores for each group (visual in C, auditory in D). Violin widths represent kernel density estimates; dashed horizontal lines indicate group means.

We next asked whether hypo- and hyper-responsivity to sensory stimuli co-occurred within the same individuals or instead reflected two distinct subgroups. Strikingly, hypo- and hyper-responsivity scores were strongly positively correlated with each other in both RTT (r=.52; P<.01) and TD (r=.74; P<.001) (Fig. 1C). These findings indicate that variability in sensory responsivity is present within individuals in both groups but is markedly elevated in RTT.

We then examined modality-specific responsivity in audition and vision: sensory modalities that can be directly and reliably probed using EP techniques.(20,21) The RTT group showed significantly higher scores than TD in both visual (Fig. 1D) and auditory (Fig. 1E) responsivity (P<.001 for both comparisons). Importantly, these modality-specific scores capture both hypo- and hyper-responsivity within each sensory domain.

These results highlight a compelling “Sensory Paradox,” in RTT, where a single individual can exhibit both hypo- and hyper-responsivity to sensory stimuli, suggesting fluctuations in responsivity rather than a shift toward a fixed phenotype.(18) We therefore next asked whether neural response variability, as measured by ITPC, could help explain this Sensory Paradox.

### EP amplitudes are dampened in RTT

We first replicated previous findings of altered EPs in RTT by examining amplitude in visual and auditory evoked potentials.

We plotted the grand-average VEP waveform at occipital electrode Oz in the standard time domain as a function of voltage (Fig. 2A), with scalp topographies confirming that Oz captured the peak visual response (Fig. 2B-C). We then extracted the mean amplitude within predefined time windows for each component for each participant. Individuals with RTT exhibited significantly reduced VEP amplitude compared to TD in both the early negative component (N1: P<.05, Fig. 2D) and the subsequent positive component (P1: P<.01, Fig. 2E). The grand-average AEP waveform was similarly examined at fronto-central electrode Fz (Fig. 2F), with scalp topographies confirming that Fz captured the peak auditory response (Fig. 2G-H). Although group differences were visually apparent, differences in AEP P1 (Fig. 2I) and N1 (Fig. 2J) amplitude did not reach statistical significance.

**Figure 2.**
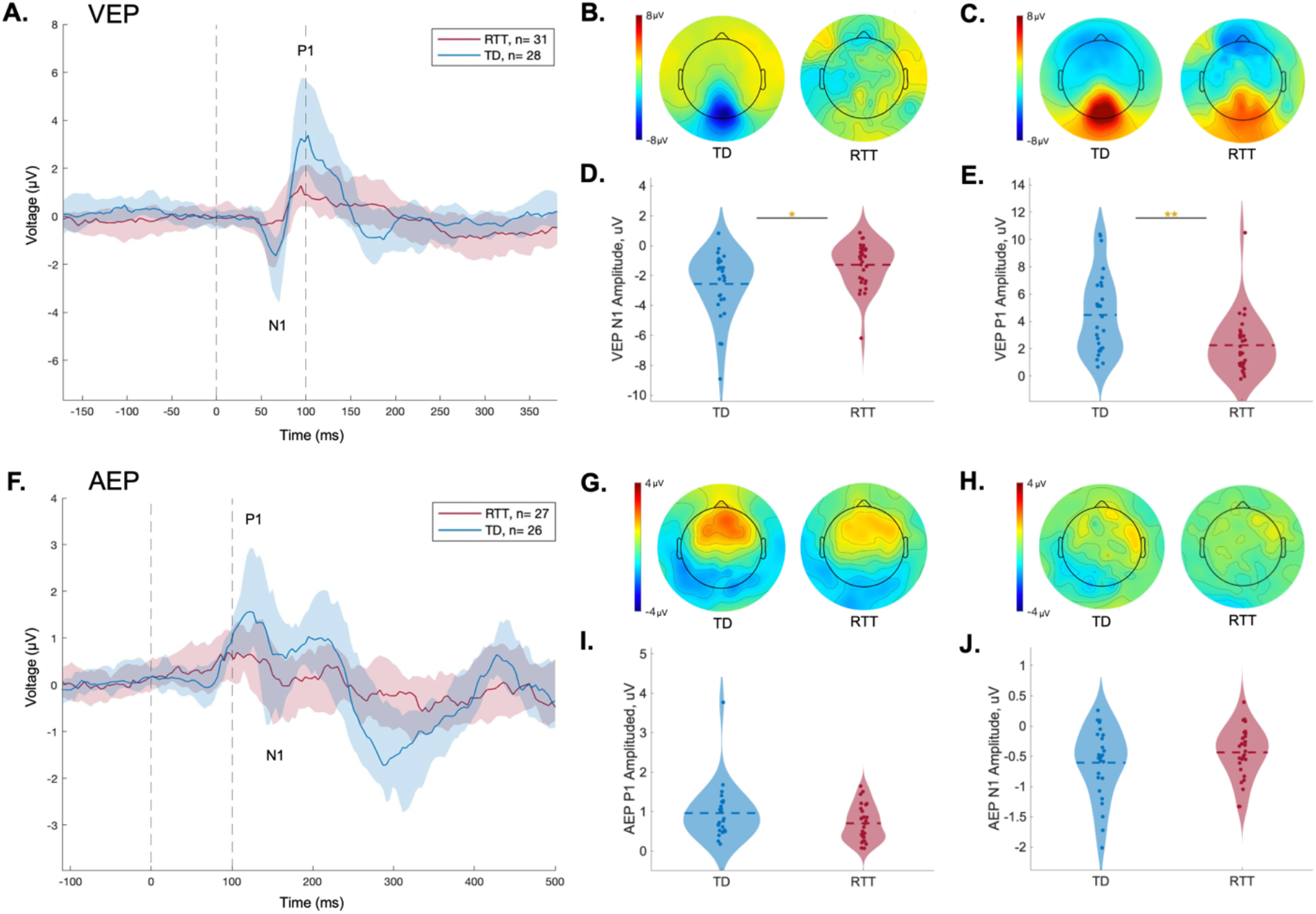
Diminished EP amplitudes in RTT. EPs in response to visual and auditory stimuli are shown for RTT (red) and TD (blue) participants. For VEP panels (A–E), RTT n=31 and TD n=28; for AEP panels (F-J), RTT n=27 and TD n=26. Statistical significance is indicated as follows: *** indicates P<0.001, ** indicates P<0.01, and * indicates P<0.05. **(A)** Grand average VEP waveforms, analyzed at occipital electrode Oz. N1 and P1 components are indicated. Shaded areas represent the 25th to 75th percentile range across participants. Vertical dashed lines indicate stimulus onset (0ms) and 100ms post-stimulus. **(B, C)** Topographic color maps display VEP voltage in all 128 electrodes at each component. **(D, E)** Violin plots of corresponding VEP N1 and P1 amplitudes at Oz, where circles represent individual participants. Violin widths represent kernel density estimates; dashed horizontal lines indicate group means. **(F-J)** All figures and analyses are repeated for the P1 and N1 components of the AEP at frontal-central electrode Fz.

It has been well established that the VEP matures early in postnatal development,(72) whereas AEP morphology continues to change across childhood and into adolescence in both TD(73,74) and RTT,(21,38). Age-stratified analyses revealed significant AEP amplitude differences in participants older than 10 years (Fig. S1A-D). Linear regression analyses confirmed age-related effects on EP amplitude: VEP P1 amplitude declined with age in both RTT (r=−.42, P<.05) and TD (r=−.41, P<.05) (Fig. S2A-B). AEP P1 amplitude also declined with age in RTT (r=−.46, P<.05) (Fig. S2C), while AEP N1 amplitude increased with age in TD (r=−.58, P<.01) (Fig. S2D), consistent with prior literature.(20,21,39) To preserve statistical power while accounting for developmental effects, all participants were included in primary analysis and age was included as a covariant where appropriate.(38)

### Inter-trial phase coherence is lower in RTT and correlates with EP amplitude

To test whether variability in neural response timing contributes to atypical sensory processing and altered EPs in individuals with RTT, we next examined ITPC in both visual and auditory evoked potentials.

Group-averaged ITPC in TD participants revealed a robust increase in ITPC following stimulus onset before returning to baseline, reflecting consistent phase-locked responses. In contrast, RTT participants showed markedly attenuated ITPC in both modalities and across frequencies (Fig. 3A,4A).

**Figure 3.**
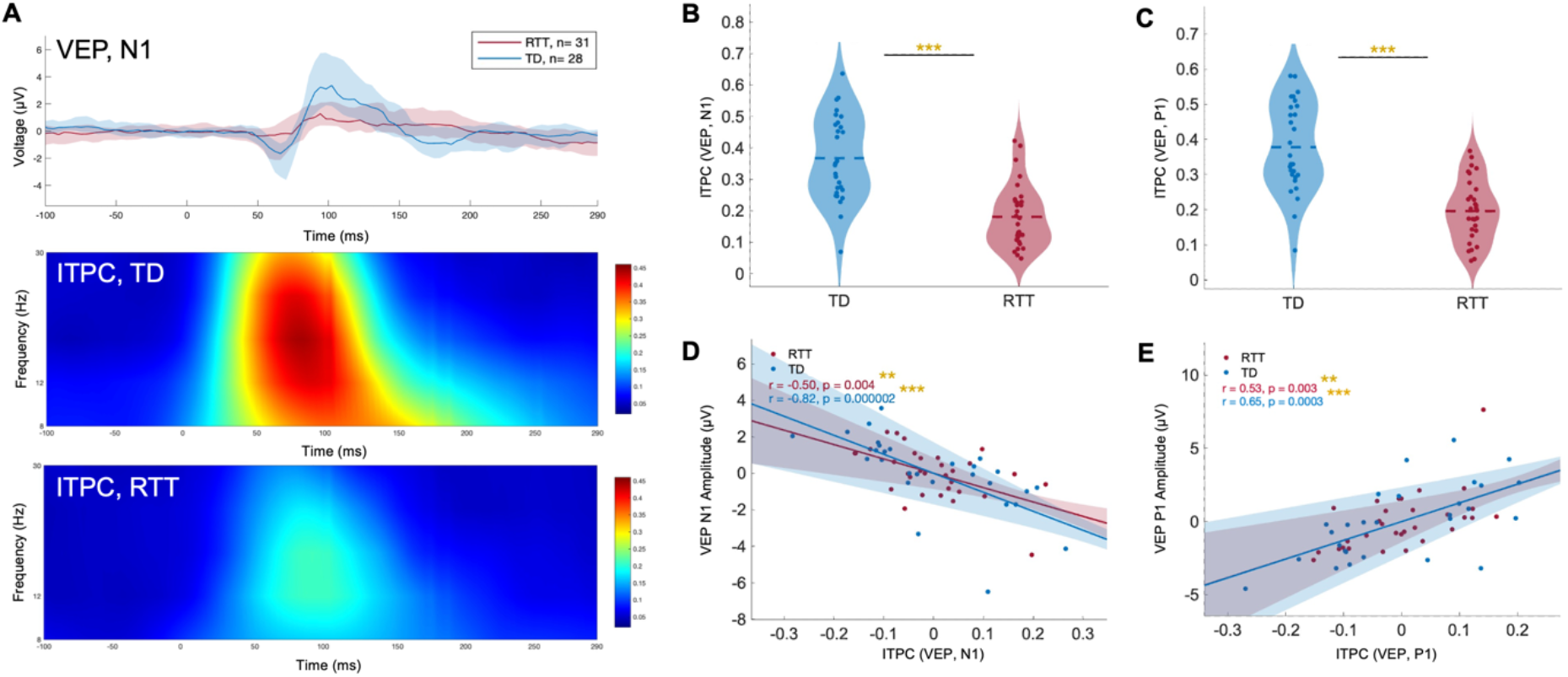
Reduced ITPC in RTT: visual-evoked potentials. EPs were analyzed at occipital electrode Oz. RTT participants are shown in red (n=31), TD participants in blue (n=28). Statistical significance is indicated as follows: *** indicates P<0.001, ** indicates P<0.01, and * indicates P<0.05. **(A)** (Top) Grand average VEP waveforms for each group; blue and red shaded areas represent the 25th to 75th percentile range across participants. Corresponding time–frequency plot shows group-averaged ITPC values for TD (middle) and RTT (bottom). **(B)** Violin plot comparing corresponding ITPC in the VEP N1 between groups, where circles represent individual participants. ITPC for each participant was averaged across the shaded N1 time window and 8–30 Hz frequencies. Violin widths reflect kernel density estimates; horizontal dashed lines indicate group mean. **(C)** Repeated for the P1 component of the VEP. **(D)** Association between VEP N1 ITPC and VEP N1 amplitude for each group. Shaded areas indicate 95% confidence intervals around linear regression lines; age was included as a covariate. (**E)** Repeated for the P1 component of the VEP.

We quantified ITPC for each participant within the same time windows used for amplitude analysis, averaging across frequencies, and assessed associations with EP amplitude using linear regression with age as a covariate. In the VEP, RTT participants exhibited significantly reduced ITPC compared to TD in both the N1 (P<.001; Fig. 3B) and P1 (P<.001; Fig. 3C) components. ITPC was strongly associated with VEP amplitude in both components and groups (RTT: N1, r=−.50, P<.01; P1, r=.53, P<.01; TD: N1, r=−.82, P<.001, P1, r=.65, P<.001, Fig. 3D-E). In the AEP, ITPC was also significantly reduced in RTT in both the P1 (P<.01, Fig. 4B) and N1 (P<.01, Fig. 4C). ITPC was associated with AEP P1 amplitude in the RTT group (r=.67, P<.001) but not TD (Fig. 4D) and was not correlated with AEP N1 amplitude in either group (Fig. 4E). Interestingly, we did not detect any association between ITPC P1 in the visual and auditory domains (Fig. S3).

**Figure 4.**
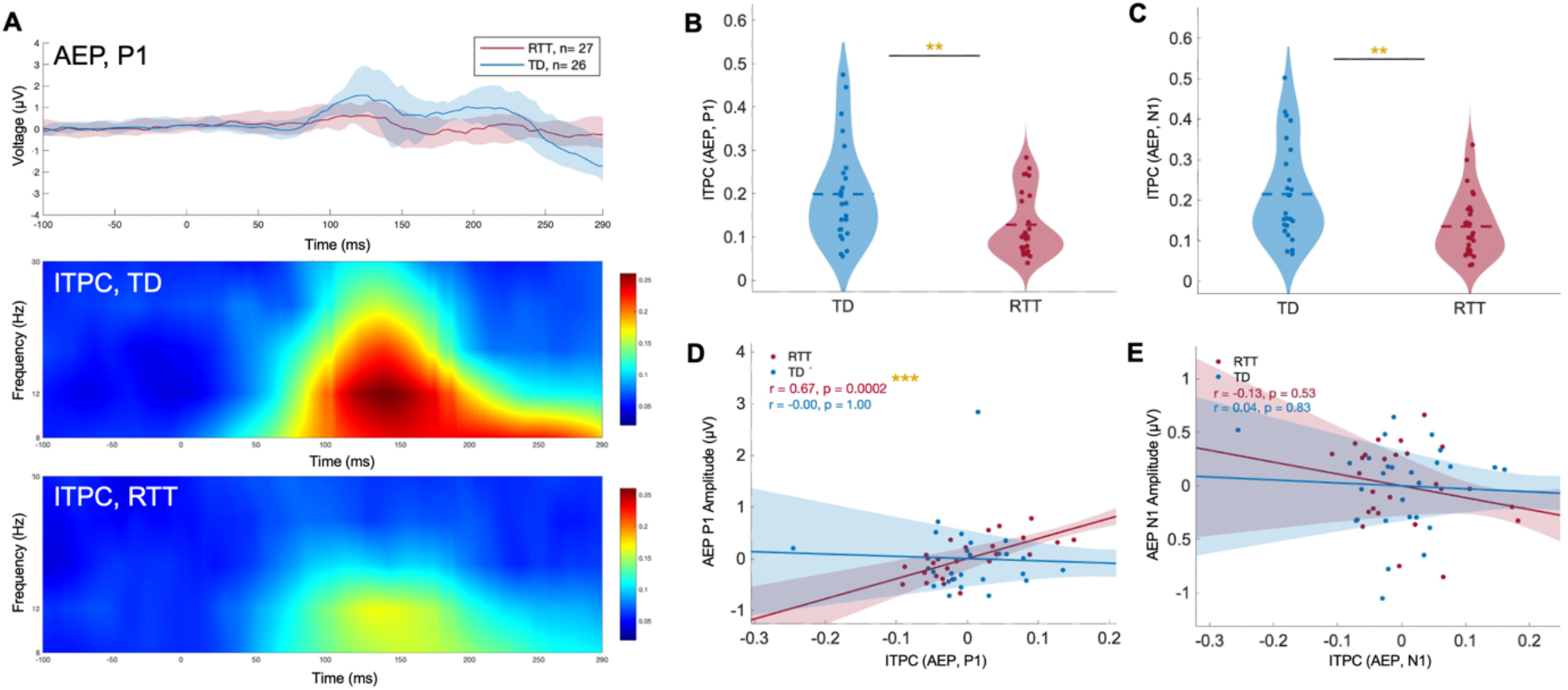
Reduced ITPC in RTT: auditory-evoked potentials. EPs were analyzed at frontal-central electrode Fz. RTT participants are shown in red (n=27), TD participants in blue (n=26). Statistical significance is indicated as follows: *** indicates P<0.001, ** indicates P<0.01, and * indicates P<0.05. **(A)** (Top) Grand average AEP waveforms for each group; blue and red shaded areas represent the 25th to 75th percentile range across participants. Corresponding time–frequency plot shows group-averaged ITPC values for TD (middle) and RTT (bottom). **(B)** Violin plot comparing ITPC in the AEP P1 between groups, where circles represent individual participants. ITPC for each participant was averaged across the shaded P1 time window and 8–30 Hz frequencies. Violin widths reflect kernel density estimates; horizontal dashed lines indicate group mean. **(C)** Repeated for the N1 component of the AEP. **(D)** Association between AEP P1 ITPC and AEP P1 amplitude for each group. Shaded areas indicate 95% confidence intervals around linear regression lines; age was included as a covariate. (**E)** Repeated for the N1 component of the AEP.

Together these findings indicate that reduced temporal alignment of neural responses across trials, as measured by ITPC, may contribute to the diminished EP amplitudes observed in RTT.

### ITPC in visual-evoked potentials is associated with sensory responsivity and behavioral symptom severity

We next examined whether ITPC was associated with sensory responsivity as measured by the SP2. Indeed, lower ITPC in both VEP components was associated with higher visual responsivity scores in RTT (N1: r=−.42, P<.05; P1: r=−.43, P<.05; Fig. 5A-B). In contrast, ITPC in AEP components was not associated with auditory responsivity (Fig. 5C-D) (even when analysis accounted for age). These results suggest a modality-specific relationship between neural response timing and behavioral sensory responsivity in RTT.

**Figure 5.**
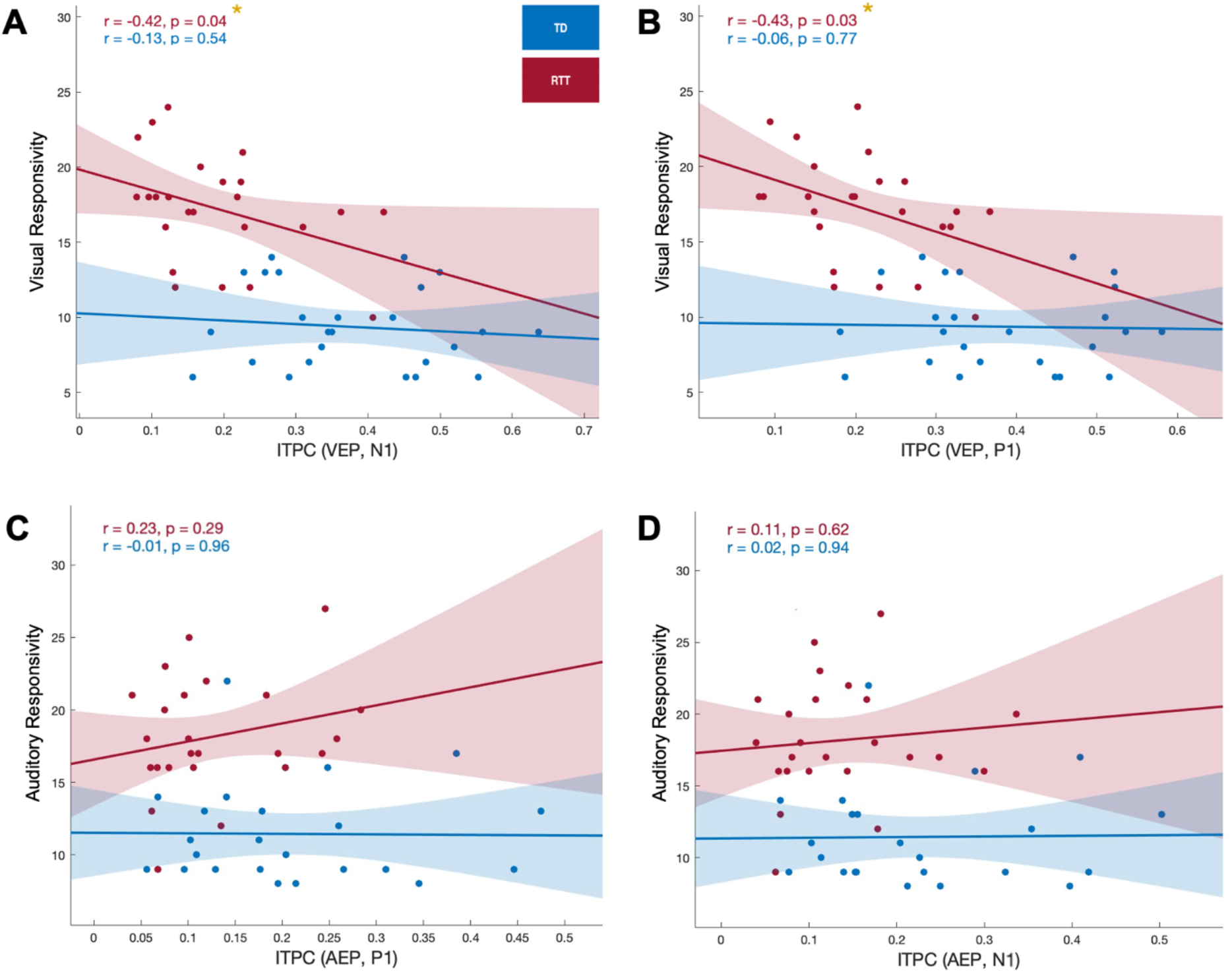
VEP ITPC is associated with visual responsivity in RTT. Association between ITPC in VEP components and visual responsivity (RTT n=24, TD n=25) **(A-B)**, and between ITPC in AEP components and auditory responsivity (RTT n=24, TD n=23) **(C-D)**. Shaded areas indicate 95% confidence intervals around linear regression lines. Statistical significance is indicated with r and P values, where: *** indicates P<0.001, ** indicates P<0.01, and * indicates P<0.05.

We then assessed the relationship between ITPC and broader clinical measures. ITPC was not associated with clinical severity (CSS and CGI-S, clinician-reported). However, lower ITPC in RTT was significantly associated with higher behavioral symptom severity (RSBQ, parent-reported) in VEP components (N1: r=−.37, P<.05; P1: r=−.37, P<.05; Fig. S4A-B), but not in AEP components (Fig. S4C-D).

ITPC was not associated with age in VEP components (Fig. S5A–B), but increased with age in the AEP P1 in both groups (RTT: r=.49, P<.01; TD: r=.79, P<.001) and in the AEP N1 in TD (r=.75, P<.001), consistent with developmental changes in auditory processing (Fig. S5C-D). RTT diagnosis subtype (typical vs atypical) and seizure frequency were not associated with ITPC.

These results indicate that reduced temporal precision, particularly in the visual domain, is associated with behavioral measures of atypical sensory responsivity and broader behavioral outcomes in RTT.

## Discussion

Atypical responsivity to sensory stimuli is a key feature of many NDDs including RTT, with important consequences for cognitive functioning, social behavior, and overall quality of life.(10– 12,75) Here, we show that both hypo- and hyper-responsivity to sensory stimuli are significantly elevated in RTT and co-occur within individuals, a Sensory Paradox consistent with increased variability in sensory processing rather than stable sensory profiles.(18) A similar, though less pronounced pattern was also observed in typically developing participants, suggesting that within-individual variability is a general property of sensory perception that is markedly amplified in RTT. At the neural level, ITPC was significantly reduced in RTT across both visual and auditory modalities and was strongly associated with diminished EP amplitudes. In the visual domain, reduced ITPC also correlated with sensory responsivity and behavioral symptom severity. These converging behavioral and electrophysiological findings support a model in which increased temporal variability in trial-by-trial neural responses contributes to both reduced EP amplitude and atypical sensory responsivity in RTT.

Neural responses to identical stimuli can vary across trials due to fluctuations in arousal, attention, and environmental context, shaping how the brain filters, amplifies, or dampens sensory information on a moment-to-moment basis. At the circuit level, such variability in RTT likely emerges from disruptions in excitatory and inhibitory (E/I) balance, a hallmark of many NDDs. *MECP2* mutations alter synaptic transmission and plasticity, leading to widespread changes in cortical network dynamics, and critically, E/I balance is not static but state-dependent and dynamically regulated.(76) RTT also involves disruptions in the relative functions of distinct inhibitory interneuron populations (“I/I” balance). GABAergic interneurons are particularly vulnerable to *Mecp2* loss(77,78), and parvalbumin-positive (PV+) cells in visual cortex show increased connectivity and activity along with enhanced thalamic inputs in *Mecp2*-deficient mice.(79,80) Supporting a key role for inhibitory circuits, selective deletion of *Mecp2* in forebrain GABAergic neurons is sufficient to produce AEP deficits, while restoring *Mecp2* expression to PV+ or somatostatin-positive (SOM+) interneurons rescues these responses.(81) VIP+ interneurons have also been implicated in RTT, with VIP-specific *Mecp2* deletion replicating key neural and behavioral phenotypes,(77) and computational modeling suggesting that reduced VIP+ excitability may underlie altered phase-amplitude coupling.(82) Although increased inhibitory tone and reduced excitatory pyramidal neuron activity is well documented, the direct contribution of these circuit dynamics to reduced ITPC remains unclear. Future studies using cell-type-specific manipulations or pharmacological modulation of inhibitory signaling will be essential to determine whether restoring E/I or I/I balance is sufficient to normalize phase coherence.

We found evidence of a modality-specific link between trial-by-trial neural variability and behavior, with reduced ITPC associated with sensory responsivity only in the visual domain, suggesting that visual cortical circuits may be especially vulnerable to loss of *MECP2*. This interpretation is supported by preclinical evidence showing visual cortical dysfunction, reduced activity, and impaired critical period plasticity.(41,42) Recent work further indicates a progressive disruption along the visual pathway: dysfunction in primary visual cortex (V1) emerges early, preceding and contributing to later alterations in thalamocortical organization, which can be prevented by restoring *Mecp2* expression.(43) These findings support a model where early cortical instability and subsequent thalamocortical degradation could contribute to more variable neural responses. Importantly, reduced ITPC likely reflects systems-level dysfunction—consistent with effects observed across the entire evoked response window rather than being confined to a single component—reflecting disruption at multiple stages of visual processing. However, it is not known where this variability originates (e.g. altered thalamocortical input, disrupted intracortical synchronization, or changes in global brain state).

Developmental factors further contribute to neural response variability, with ITPC increasing with age in AEP components. Longitudinal studies will be necessary to determine how ITPC evolves over the course of the disorder and whether deficits emerge early or progress over time; such work would help establish ITPC as a functional biomarker capable of tracking disease progression and treatment efficacy.

Taken altogether, our data demonstrate a Sensory Paradox in RTT and identify reduced consistency in trial-by-trial neural responses, measured by ITPC, as a potential mechanism linking atypical sensory responsivity to altered cortical processing. These findings support ITPC as a translational biomarker, and given that atypical sensory processing is a common feature of many NDDs (including both idiopathic and syndromic ASDs), may represent a shared pathophysiological mechanism and a promising target for future therapeutic studies.

## Data Availability

Data is available on reasonable request.

## Author Contributions

DK, AL, and MF conceptualized the study. DK wrote and prepared the initial manuscript draft. DK and KNS collected clinical data. DK performed the analysis. DK, AL, MF, and CN interpreted the analytic findings. All authors contributed to interpretation of the results and critical revision of the manuscript.

## Acknowledgements

Thank you very much to all the participants in these studies and their families, especially those who travelled and extended themselves to contribute to research on Rett syndrome. Thank you to the Rett Syndrome Angels (originally Rett Syndrome Association of Massachusetts) for their incredible support. Thank you to the Rett syndrome clinics and to all members of the labs of Dr.’s April Levin, Michela Fagiolini, Charles Nelson, and Takao Hensch for your invaluable feedback and contributions. This work could not have been completed without all of you.

## Funding

This work was supported partially by:

P50HD105351 (Project 1) and the International Research Center for Neurointelligence (IRCN) - Michela Fagiolini

