## Supplemental Figure 1 for "Variability in sensory processing and evoked potentials in Rett syndrome"

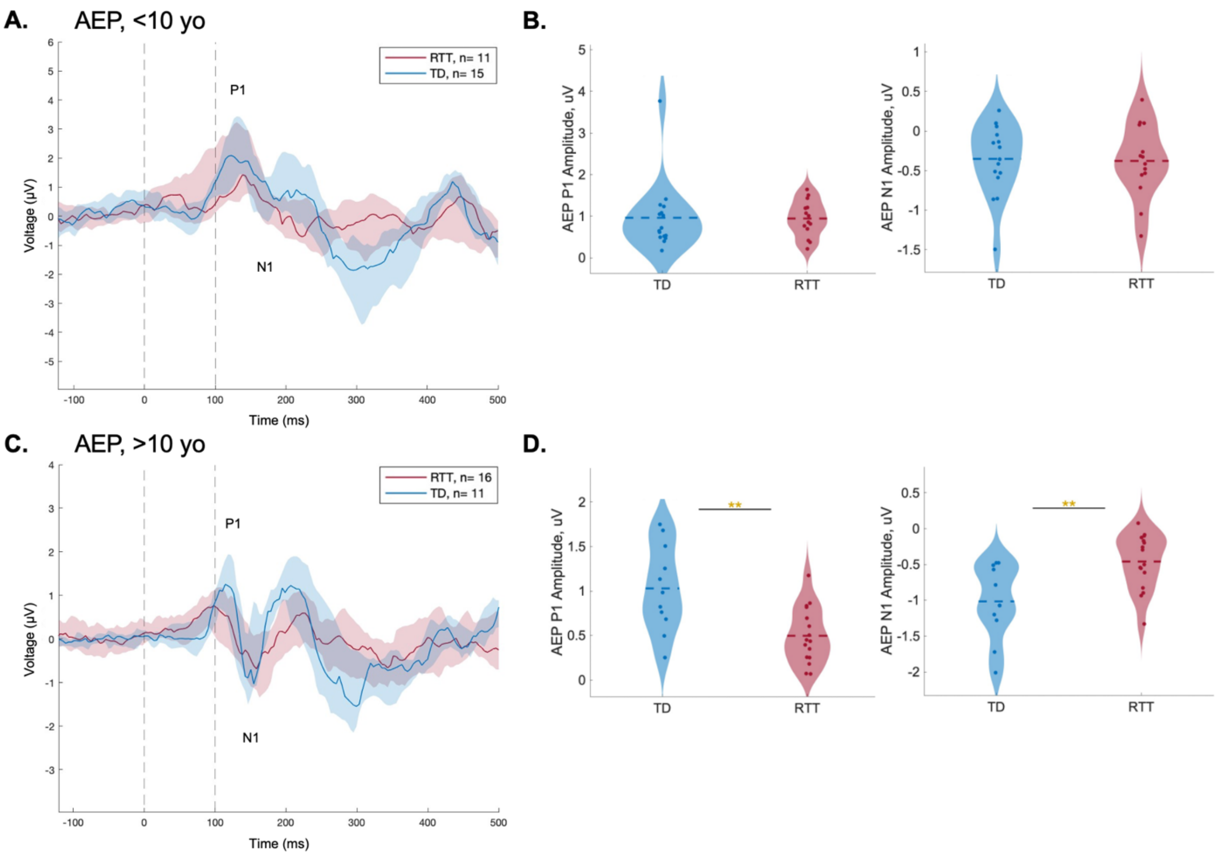


**Figure S1. Difference in AEP amplitude emerges when stratifying by age.** AEPs are shown for RTT (red) and TD (blue) participants in two age groups. **(A)** Grand average AEP waveforms are shown for participants below 10 years old (RTT n=11 and TD n=15). The P1 and N1 components are indicated. AEP was analyzed at frontal-central electrode Fz. Shaded areas represent the 25th to 75th percentile range across participants. Vertical dashed lines indicate stimulus onset (0ms) and 100ms post-stimulus. **(B)** Violin plots of corresponding AEP P1 and N1 amplitudes, where circles represent individual participants. Statistical significance is indicated, where *** indicates P<0.001, ** indicates P<0.01, and * indicates P<0.05. Violin widths represent kernel density estimates; dashed horizontal lines indicate group means. **(C-D)** Figures and analyses are repeated for participants above 10 years old (RTT n=16 and TD n=11).
