## Supplemental Figure 2 for "Variability in sensory processing and evoked potentials in Rett syndrome"

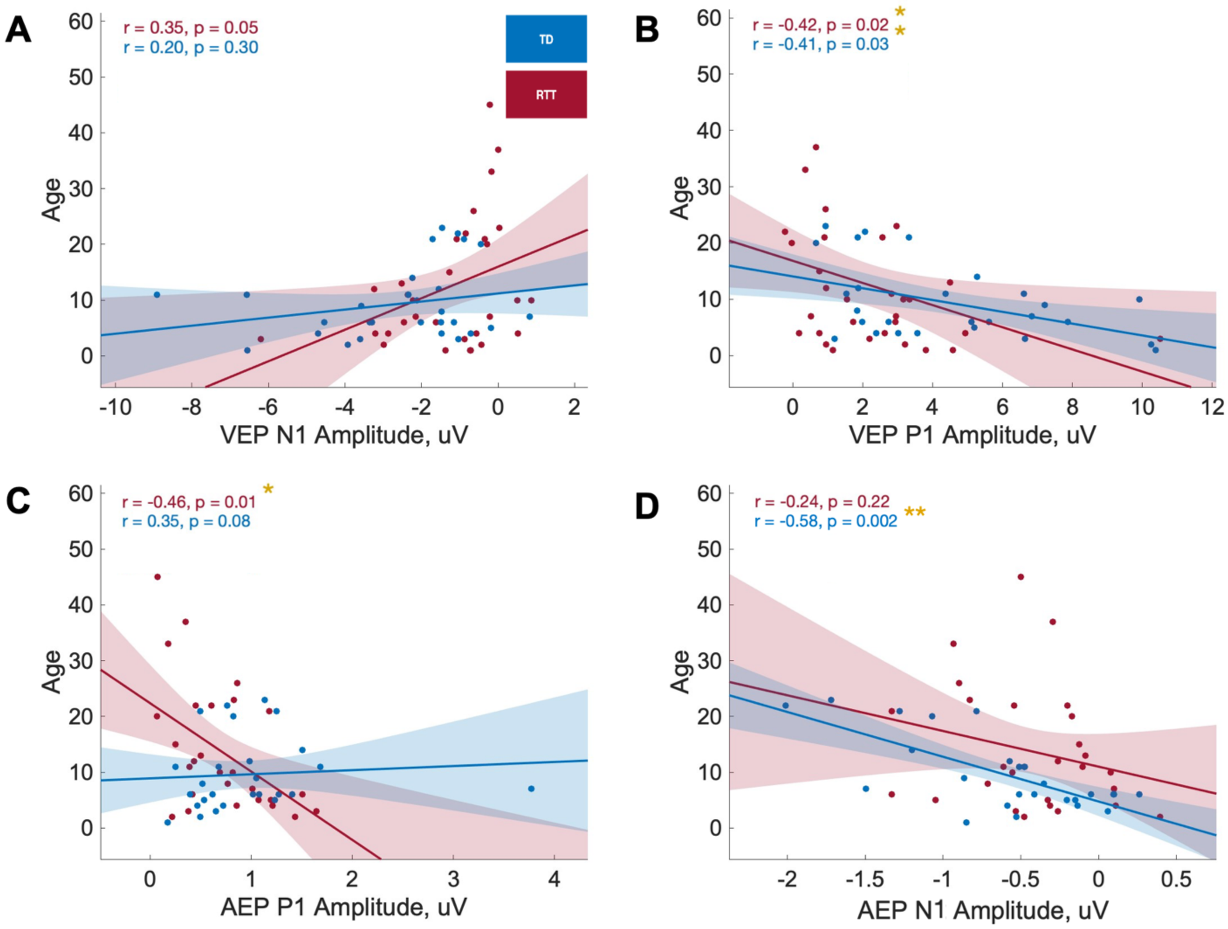


**Figure S2. Amplitude in some EP components is associated with age.** Association between amplitude in VEP components and age (RTT n=31, TD n=28) **(A-B)**, and between amplitude in AEP components and age (RTT n=27, TD n=26) **(C-D)**. Shaded areas indicate 95% confidence intervals around linear regression lines. Statistical significance is indicated with r and P values, where: *** indicates $P<0.001$, ** indicates $P<0.01$, and * indicates $P<0.05$.
