## Supplemental Figure 3 for "Variability in sensory processing and evoked potentials in Rett syndrome"

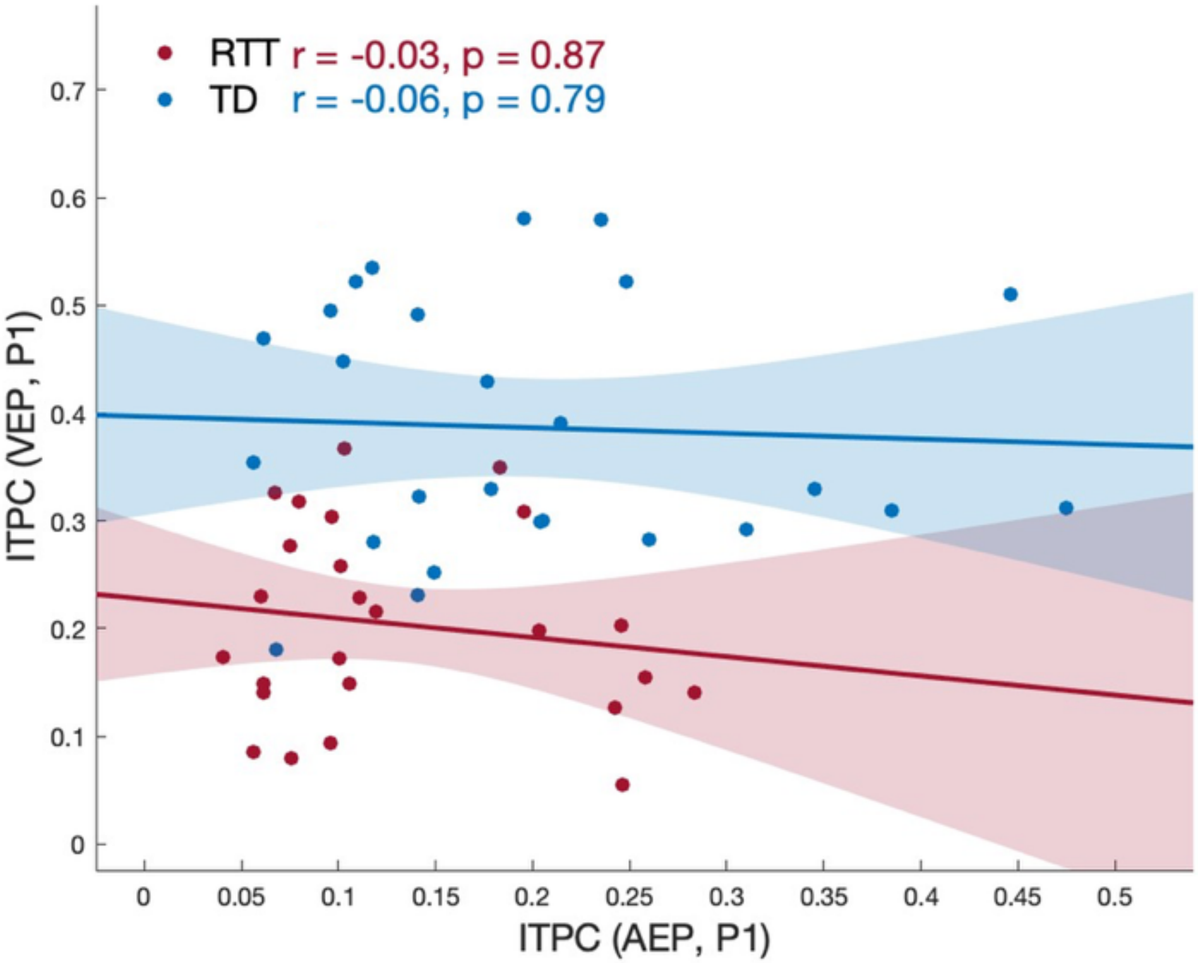


**Figure S3. ITPC in visual- and auditory-evoked responses are not associated with each other.** Association between ITPC in the VEP P1 component and ITPC in the AEP P1 component. RTT n=25 and TD n=26. Shaded areas indicate 95% confidence intervals around linear regression lines. Statistical significance is indicated with r and P values, where: *** indicates P<0.001, ** indicates P<0.01, and * indicates P<0.05.
