## Supplemental Figure 4 for "Variability in sensory processing and evoked potentials in Rett syndrome"

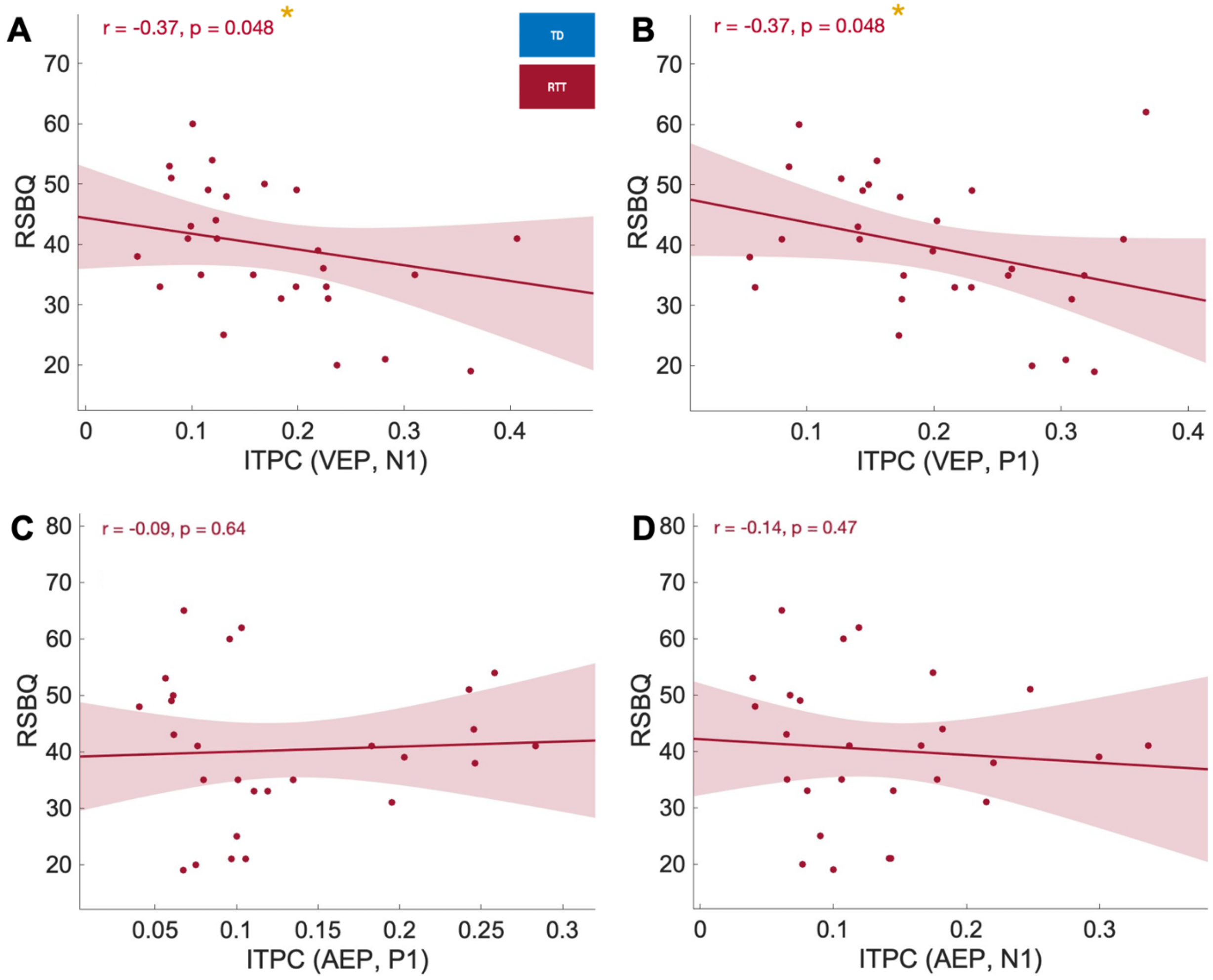


**Figure S4. VEP ITPC is associated with behavioral symptom severity in RTT.** Association between ITPC in VEP components and RSBQ score (n=29) **(A-B)**, and between ITPC in AEP components and RSBQ score (n=27) **(C-D)**. Shaded areas indicate 95% confidence intervals around linear regression lines. Statistical significance is indicated with r and P values, where: *** indicates P<0.001, ** indicates P<0.01, and * indicates P<0.05.
